# Modelling the Impact of Nationwide BCG Vaccine Recommendations on COVID-19 Transmission, Severity, and Mortality

**DOI:** 10.1101/2020.05.10.20097121

**Authors:** Nita H. Shah, Ankush H. Suthar, Moksha H. Satia, Yash Shah, Nehal Shukla, Jagdish Shukla, Dhairya Shukla

## Abstract

COVID-19 was declared as pandemic on 11^th^ March 2020 by WHO. There are apparent dissimilarities in incidence and mortality of COVID-19 cases in different parts of world. Developing countries in Asia and Africa with fragile health system have shown lower incidence and mortality compared to developed countries with superior health system in Europe and America. Most countries in Asia and Africa have national BCG vaccination program while Europe and America do not have such program, or have ceased it. At present, there is no known therapy to treat COVID-19 disease. There is no vaccine available currently to prevent COVID-19 disease. As mathematical modelling is ideal for predicting the rate of disease transmission as well as evaluating efficacy of possible public health prevention measures, we have created a mathematical model with seven compartments to understand nationwide BCG vaccine recommendation on COVID-19 transmission, severity and mortality. We have computed two basic reproduction number, one at vaccine free equilibrium point and other at non-vaccine free equilibrium point and carried out local stability, sensitivity and numerical analysis. Our result showed that individuals with BCG vaccinations have lower risk of getting COVID-19 infection, shorter hospital stays, and increased rate of recovery. Furthermore, countries with long-standing universal BCG vaccination policies have reduced incidence, mortality, and severity of COVID-19. Further research will focus on exploring the immediate benefits of vaccination to healthcare workers and patients as well as benefits of BCG re-vaccination.

## 1. Introduction

In December of 2019, a novel strain of coronavirus was found in Wuhan, China and identified as Coronavirus Disease 2019 (COVID-19) [1]. Transmission of COVID-19, similar to numerous airborne respiratory viruses including tuberculosis and influenza, occurs through direct contact with an infected person through respiratory droplets when a person coughs or sneezes or indirect contact through interaction with contaminated surfaces with respiratory droplets from infected person [2]. As of April 18, 2020, the disease has spread to 210 countries with 2,324,731 total confirmed new cases reported and 160,434 deaths [3]. Since currently there is no definitive treatment or an effective vaccine for disease control, there is a crucial need to find measures that cure or reduce morbidity due to COVID-19.

There are prominent dissimilarities in how COVID-19 is affecting different countries. Although developing countries like India, Philippines, Sri Lanka reported their initial case in January, they have yet to experience extensive community spread [5]. In contrast, developed countries like Italy and United Kingdom have experienced high mortality and widespread infection despite strong curtailing of social interactions. Studies show a disproportionately smaller number of cases have been reported from disadvantaged and low-income countries [6]. It is puzzling that countries with more fragile health systems report less incidence of COVID-19. Figure 1 shows a figure from the European Centre for Disease Prevention and Control which indicates that much of the incidence of COVID-19 is in the developing countries such as United Kingdom and United States [4]. A possible explanation for lower number of cases detected in these developing countries with extensive travel and trade links to China may stem from the induced heterologous protective activity of the Bacillus Calmette-Guerin (BCG) immunization [5].

**Figure 1:**
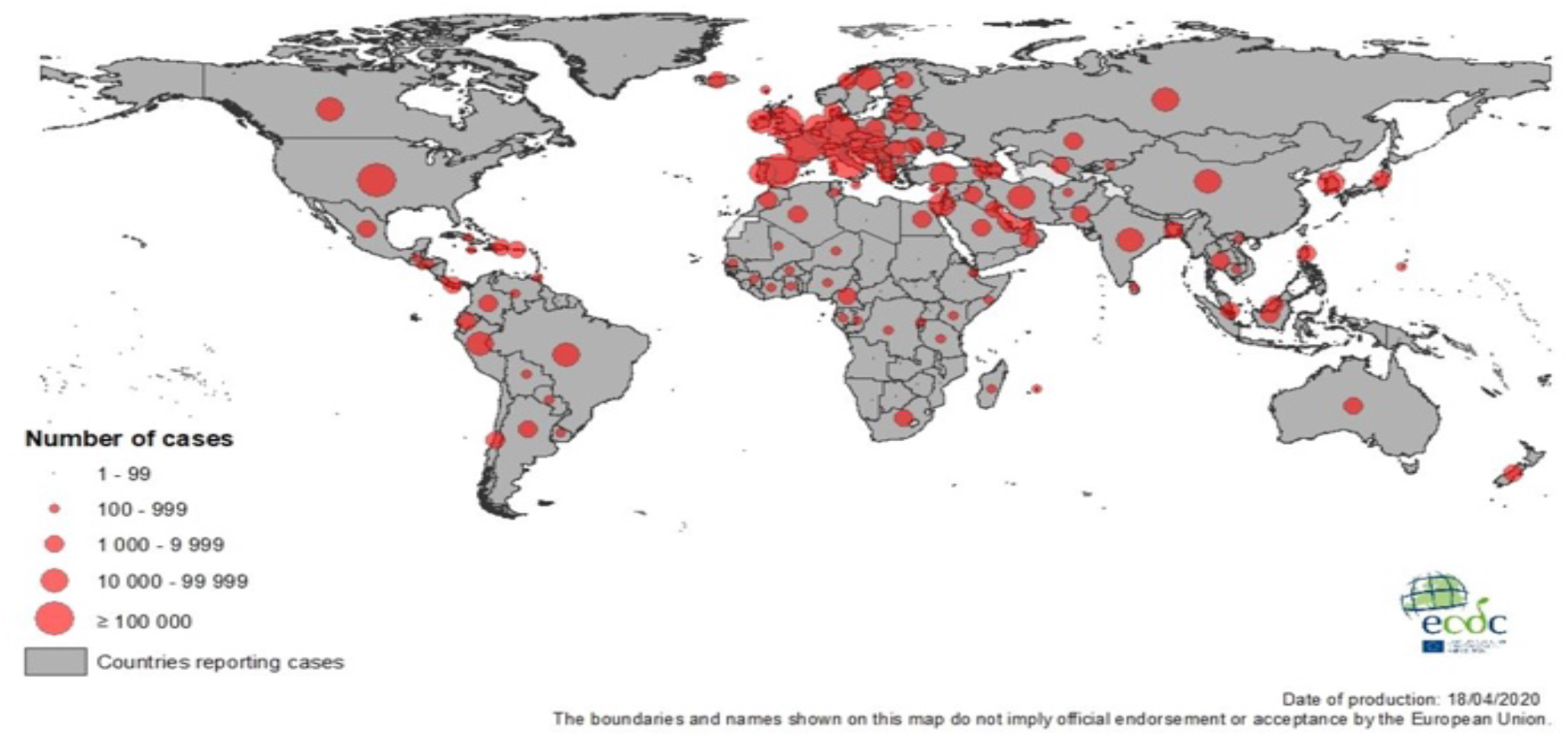
Geographical distribution of COVID-19 cases worldwide as of 18^th^ April 2020[4]. *Adapted from European Centre for Disease Prevention and Control*.

Vaccines provide protection from particular pathogens by inducing effector mechanisms directed towards that pathogen. Certain live attenuated vaccines like the BCG vaccine increase immunity against not only a particular pathogen, but also numerous unrelated pathogens that present with similar acute respiratory tract infections [8]. Previous studies have established that BCG vaccination confers non-specific protection via the induction of innate immune memory [9]. BCG is used widely across the world as a vaccine for Tuberculosis, with many developing nations having a universal BCG vaccination policy for new-born[10]. Studies have indicated that countries with uniform BCG vaccination policy show significantly lower COVID-19 cases and deaths per million people compared to countries where BCG vaccination policy was ceased or was never in place [5]. Figure-2 from the BCG world atlas shows that numerous developing countries currently employ a universal BCG vaccination program, while many developed countries have either ceased their BCG vaccination recommendation or never recommended it [7]. Comparison of Figures 1 and 2 also indicates that the largest number of COVID-19 cases are in countries that currently have no BCG vaccination recommendation.

**Figure 2:**
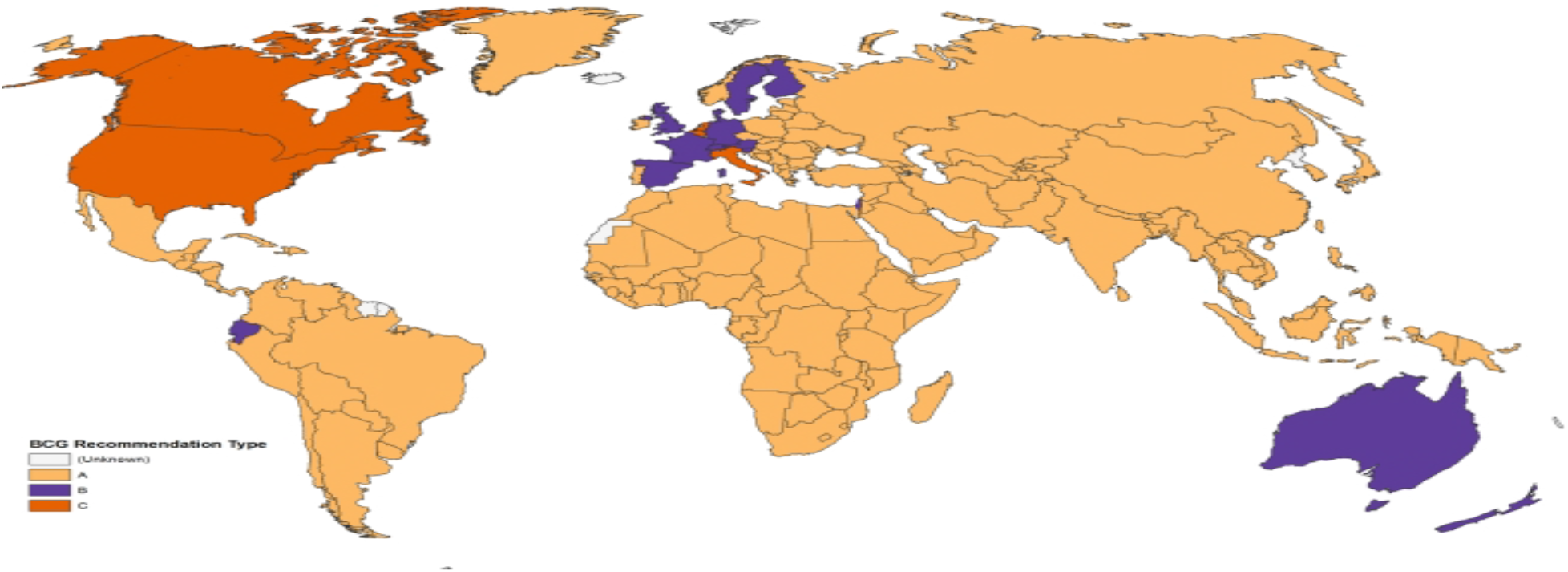
Map displaying BCG vaccination policy by country. A: The country currently has universal BCG vaccination program. B: The country used to recommend BCG vaccination for everyone, but currently does not. C: The country never had universal BCG vaccination programs [7]. *Adapted from BCG World Atlas*.

Furthermore, the year that universal BCG vaccination was established within a country has a significant correlation with the mortality rate, indicating that earlier implementation of the policy leads to protection of a larger fraction of the elderly population. For instance, although Iran currently employs a universal BCG vaccination policy, it was implemented as recently as 1984. Consequently, it reports an elevated mortality with 19.7 deaths per million inhabitants[6]. In contrast, Japan started its universal BCG policy in 1947 and has around 100 times less deaths per million people, with 0.28 deaths. Similarly, Brazil started universal vaccination in 1920 and also has an even lower mortality rate of 0.0573 deaths per million inhabitants [6].

Exploring this relationship between BCG vaccination and COVID-19 further is crucial in establishing a potential short- or long-term prevention measure that is effective in reducing incidence and mortality of COVID-19. While previous studies have explored a possible link between the BCG vaccine and incidence of COVID-19, they have not modelled the impact of implementing a nationwide BCG immunization policy [6,7]. Mathematical modelling is ideal for predicting the rate of disease transmission as well as evaluating efficacy of possible public health prevention measures [11]. This study aims to create a model to evaluate the effect of nationwide BCG vaccine recommendations on incidence, severity, and mortality rates of COVID-19. Furthermore, our study aims to model the protentional implications of an immediate implementation of the universal BCG vaccination policy.

## 2. Mathematical Mode

A novel corona virus pandemic is occurring around the world, both in countries with and without the BCG vaccine. To analyse the importance of this vaccine against the fight of COVID-19, we have developed a mathematical model. This model has seven various compartments. First compartment is the class of exposed individuals to COVID-19 noted as *E*. From these individuals, some are vaccinated with BCG vaccine noted as *V* and those who have not taken it are noted as *N_V_*. Now, with the certain rate these individuals transfer into Infected stage noted as *I* and some can become critically infected noted as *C* compartment. The individuals suffering from COVID-19 that need to be hospitalised are noted as *H*. The infected and hospitalised individual that recover are noted as *R*.

With the help of transmission shown in figure 3 and notation of parameter given in table 1, the system of non-linear ODE is derived as;

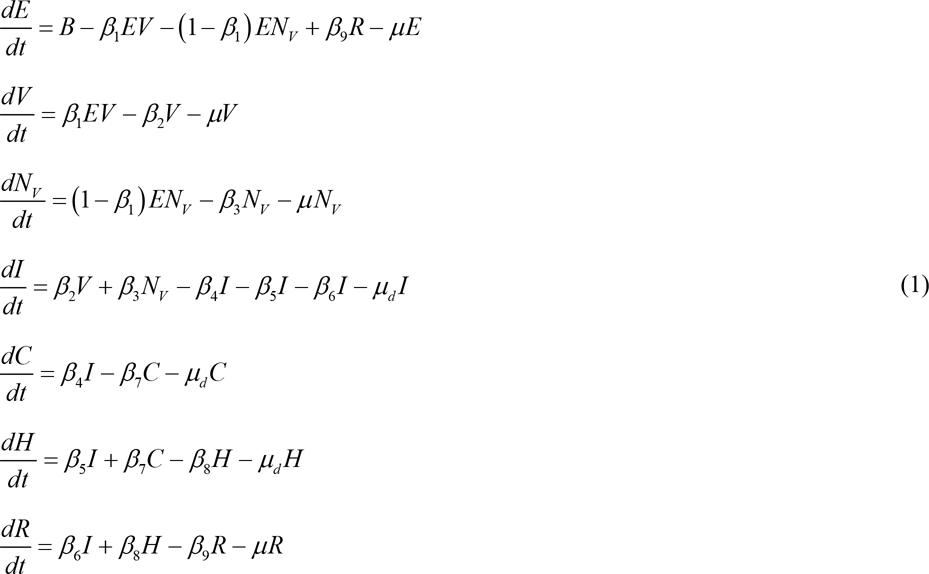

with *E* > 0; *V*, *N_V_*, *I*, *C*, *H*, *R* ≥ 0

**Table 1:** Notation and its description.

| Parameter | Description |
| --- | --- |
| $B$ | Birth rate of class of exposed individuals |
| $\beta_1$ | Rate of individual who has taken BCG vaccine |
| $\beta_2$ | Rate of vaccinated individuals who get infection |
| $\beta_3$ | Rate of non-vaccinated individuals who get infection |
| $\beta_4$ | Rate at which infected individual become critically infected |
| $\beta_5$ | Rate at which infected individuals get hospitalised |
| $\beta_6$ | Recovery rate of infected individuals |
| $\beta_7$ | Rate at which critically infected individuals get hospitalised |
| $\beta_8$ | Recovery rate of hospitalised individuals |
| $\beta_9$ | Rate at which recovered individuals may get exposed |
| $\mu$ | Natural death rate |
| $\mu_d$ | Diseases induced death rate |

**Figure 3:**
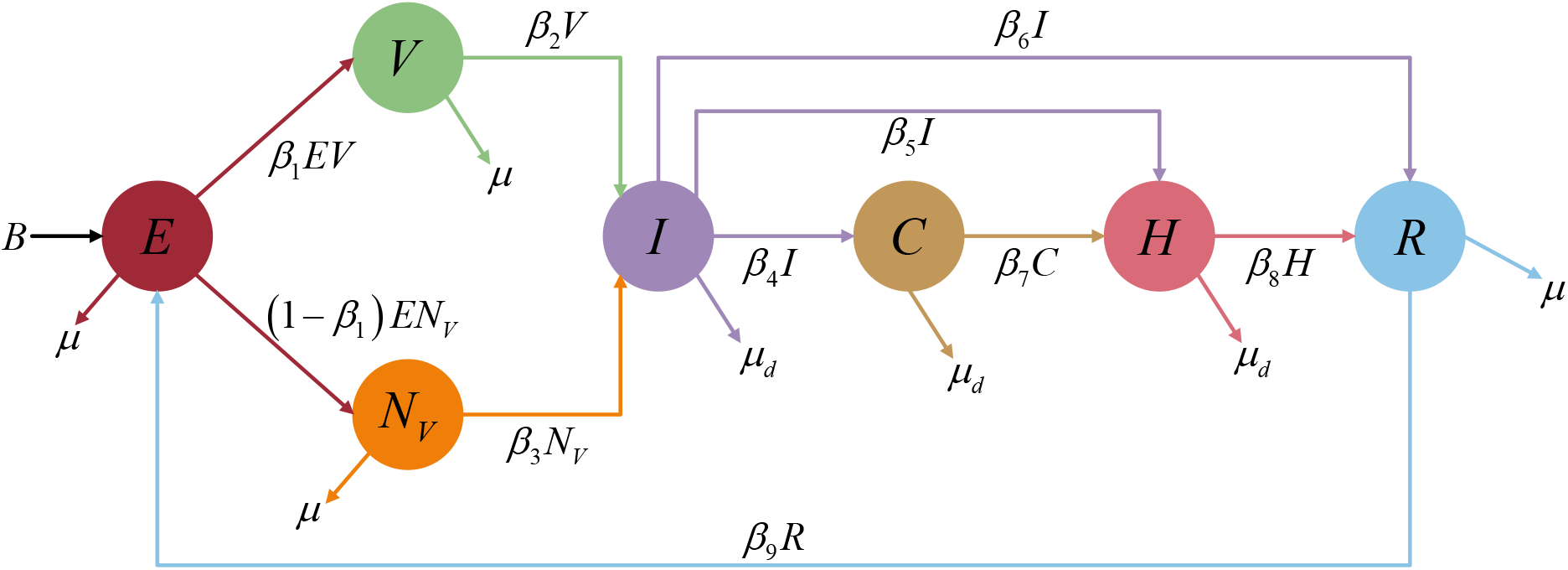
Transmission diagram of COVID-19 with BCG vaccine.

Hence, the feasible region of given system is

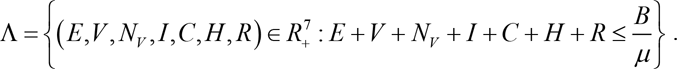

Now, consider *β*_2_ + *μ* = *k*_1_, *β*_3_ + *μ* = *k*_2_, *β*_4_ + *β*_5_ + *β*_6_ *+ μ_d_* = *k*_3_, *β*_7_ + *μ_d_* = *k*_4_, *β*_8_ + *μ_d_* = *k*_5_, *β*_9_ + *μ_d_* = *k*_6_ then we get new system as follow

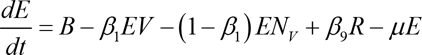

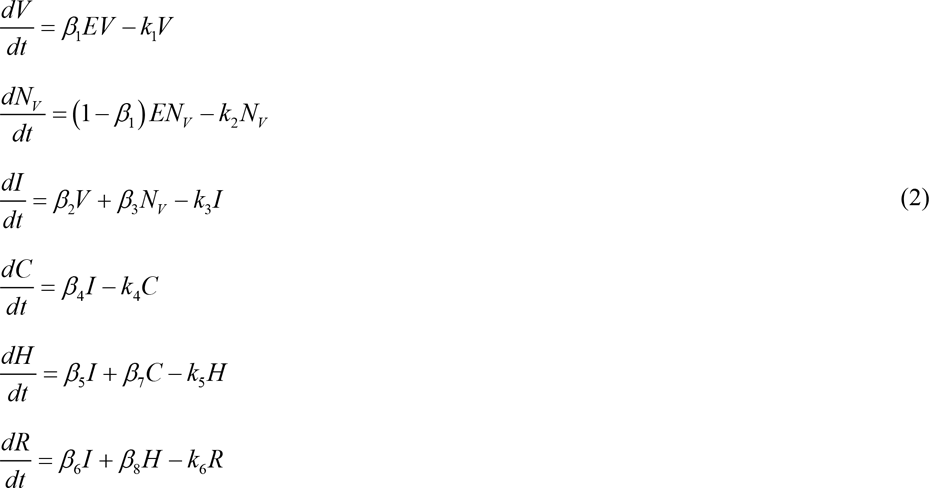

The dynamical behaviour of system (1) is equivalent to the system (2). Hence, every solution of system (2) will remain in the region Λ.

The above system (2) has three equilibrium point by setting the equation as zero.

1. Diseases-free equilibrium point 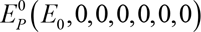 where 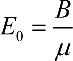
2. Vaccine free equilibrium point 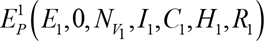 where

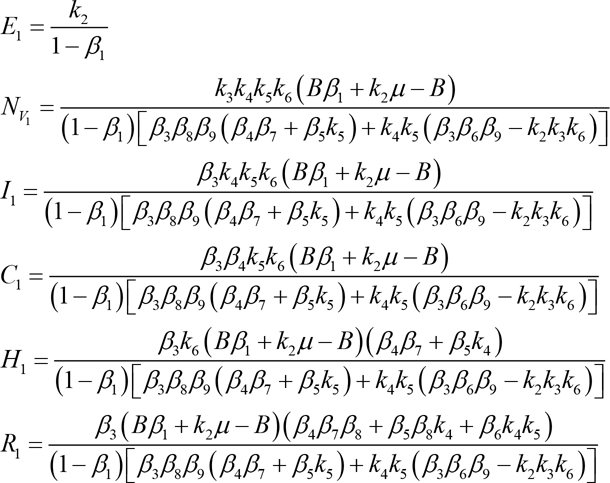
3. Non-vaccine free equilibrium point 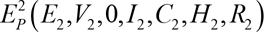

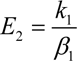

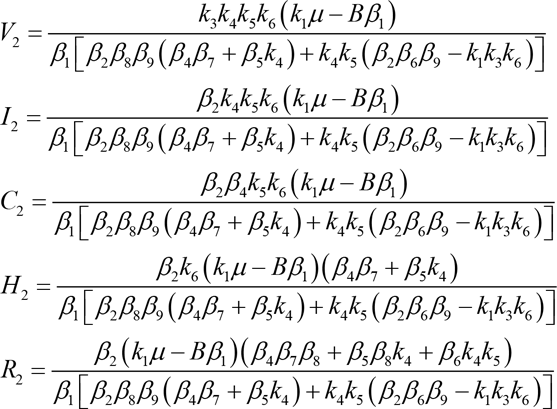 The diseases-free equilibrium point is not valid according to current scenario and therefore we will discuss only two equilibrium points.

## 3. Basic Reproduction Number

In this section, basic reproduction number is derived using next generation matrix method [12]. The quantity of basic reproduction number helps to understand the behaviour of the spread of COVID-19 amongs the polpulation. The basic reproduction number is denoted as *R*_0_. If the value of *R*_0_ is less than 1 then the diseases is in the controlable stage. Otherwise, it has reached the epidemic stage. This is the ratio of affected infected individual by secondary infected individuals within the population. Using next generation matrix method, *f* and *v* are derived as

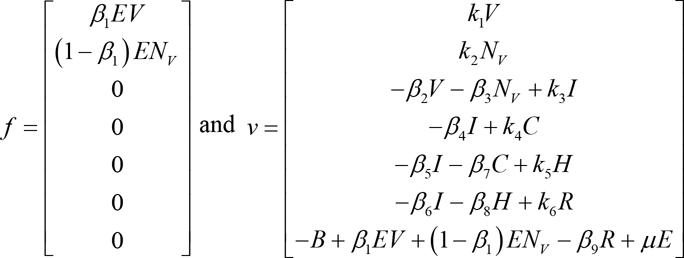

The Jacobian matrices of *f* and *v* are *F* and *V* defined as below

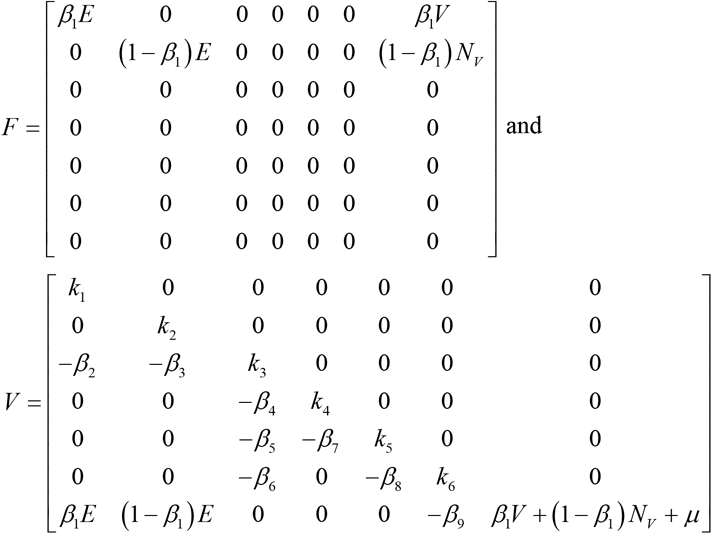

These Jacobian matrices are used to derive *FV^−^*^1^ whose spectral radius is our desired basic reproduction number (*R*_0_). Here, we have computed two basic reproduction numbers; one at vaccine free equilibrium point and other at non-vaccine free equilibrium point.

The expression of basic reproduction number for non-vaccinated individuals i.e. at vaccine-free equilibrium point is denoted by 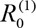 given by

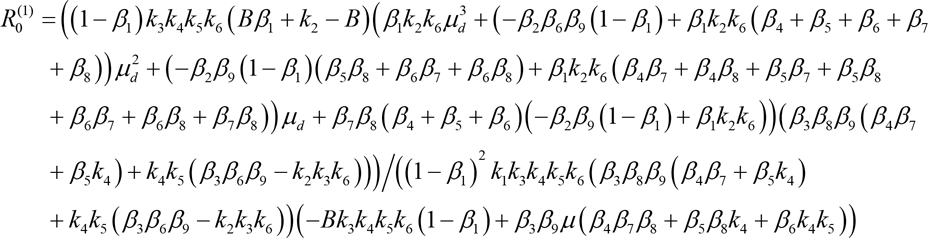

Whereas, the expression of basic reproduction number for vaccinated individuals i.e. at nonvaccine-free equilibrium point is denoted by 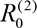 given by

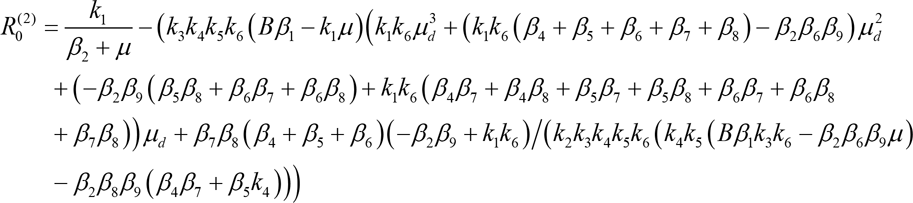

Moreover, the Basic reproduction for the system is 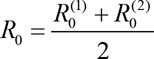

## 4. Local Stability Analysis

In this section, we will be discussing the local stability for vaccine and non-vaccine free equilibrium point utilizing the concept of eigenvalues. To find the eigen values, the Jacobian matrix of the system (2) is calculated as

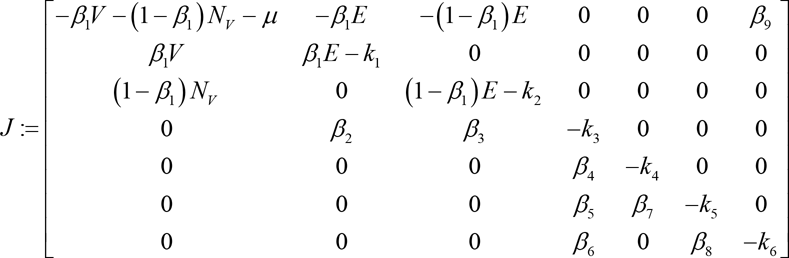

Eigenvalues for above Jacobi matrix about the vaccine-free equilibrium point are 0.1225, - 0.0664, −0.1375, −0.1375, −0.8463, −0.9431, 0.7019.

Eigenvalues for above Jacobi matrix about the non-vaccine-free equilibrium point are 0.2298, −0.0655, −0.1595, −0.2296, −0.8464, −0.9431, −0.1021.

Hence, both the equilibrium points are not locally asymptotically stable.

## 5. Sensitivity Analysis

Here, sensitivity analysis is carried out to see which parameter is conferring a positive effect on the model. The sensitivity analysis is conducted on the basic reproduction number. It is to be done using Christoffel formula which is 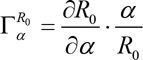 where *α* is the model parameter.

**Table 2:**
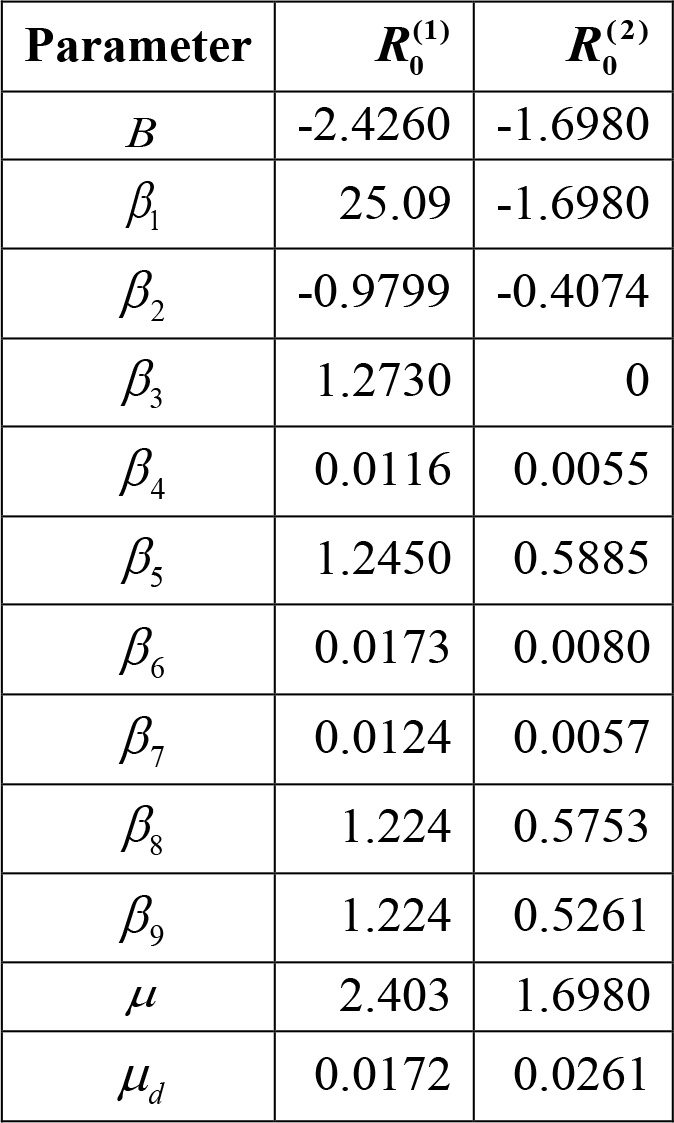
Sensitivity Analysis.

From table 2, it can be observed that the growth rate is producing negative impact. Precisely, growth rate is more negative when there is no vaccinated individual. The Rate of individual vaccinated with BCG (*β*_1_) shows the positive impact among all. Moreover, the disease induced death rate is lower for 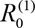 when compared to 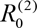. Additionally, considering the sensitivity analysis for both the basic reproduction numbers, we conclude that BCG vaccine is beneficial in preventing COVID-19.

## 6. Numerical Simulation

In this section, results of transmission of COVID-19 outbreak is simulated numerically, helping validate model results.

**Figure 4:**
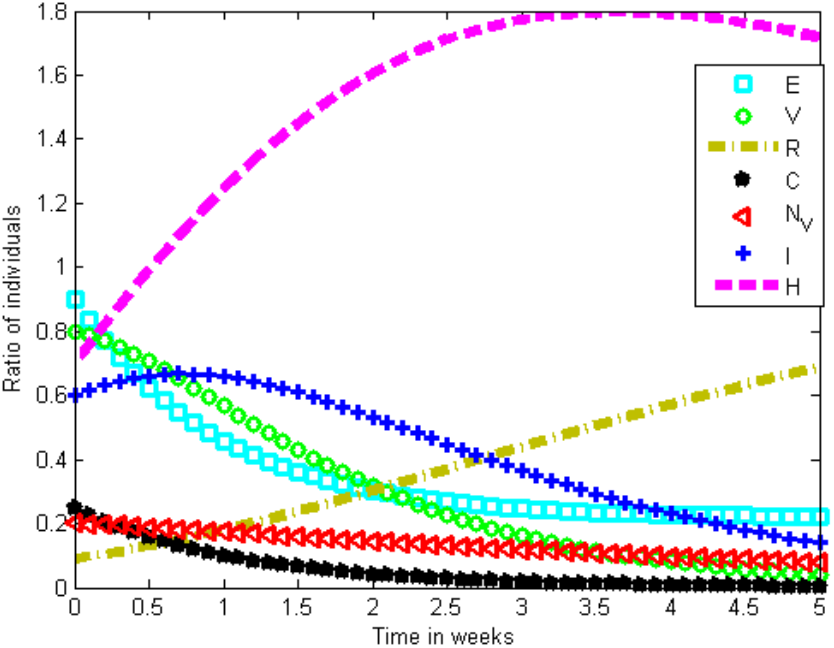
Transmission of COVID-19.

Figure 4 shows the change in the behaviour of individuals. This suggests that exposed individuals quickly progress into infected ones. Infected individuals increase in first 1.5 weeks and then start to decrease whereas critically infected individuals stay in lower numbers. Hospitalisation increases rapidly for at least 3 weeks and the it flattens. Recovery rate increases but at lower rate.

**Figure 5:**
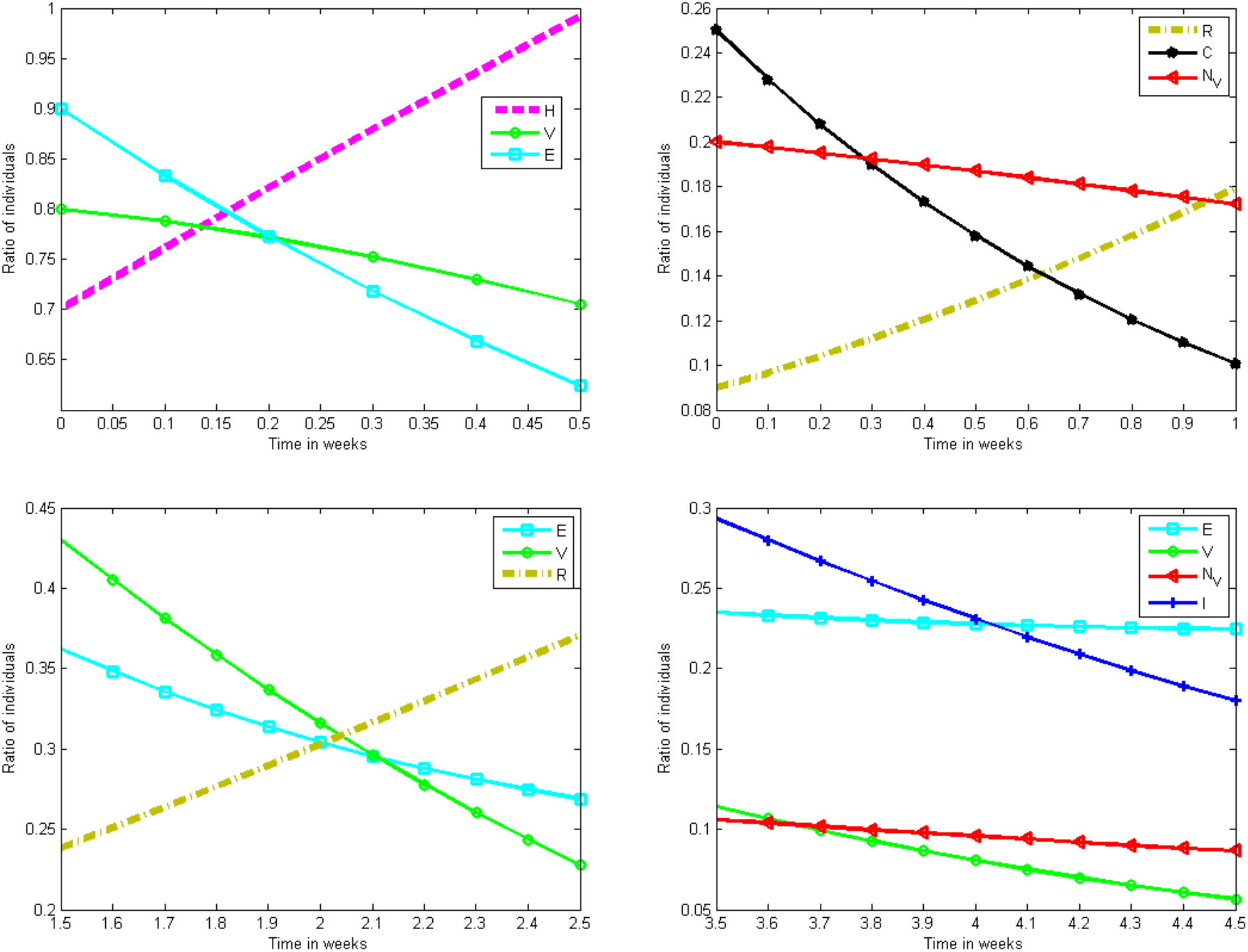
Magnified plots of transmission of COVID-19.

To get precise results, four clusters of figure 5 is magnified. Our data indicates that individuals should go into isolation earlier to prevent exposure to COVID-19. BCG vaccinated individual may get exposed in 0.2 week while, at the same time, non-vaccinated individual may get critically infected. Moreover, recovered individual also may become exposed and get re-infected after 2 weeks.

**Figure 6:**
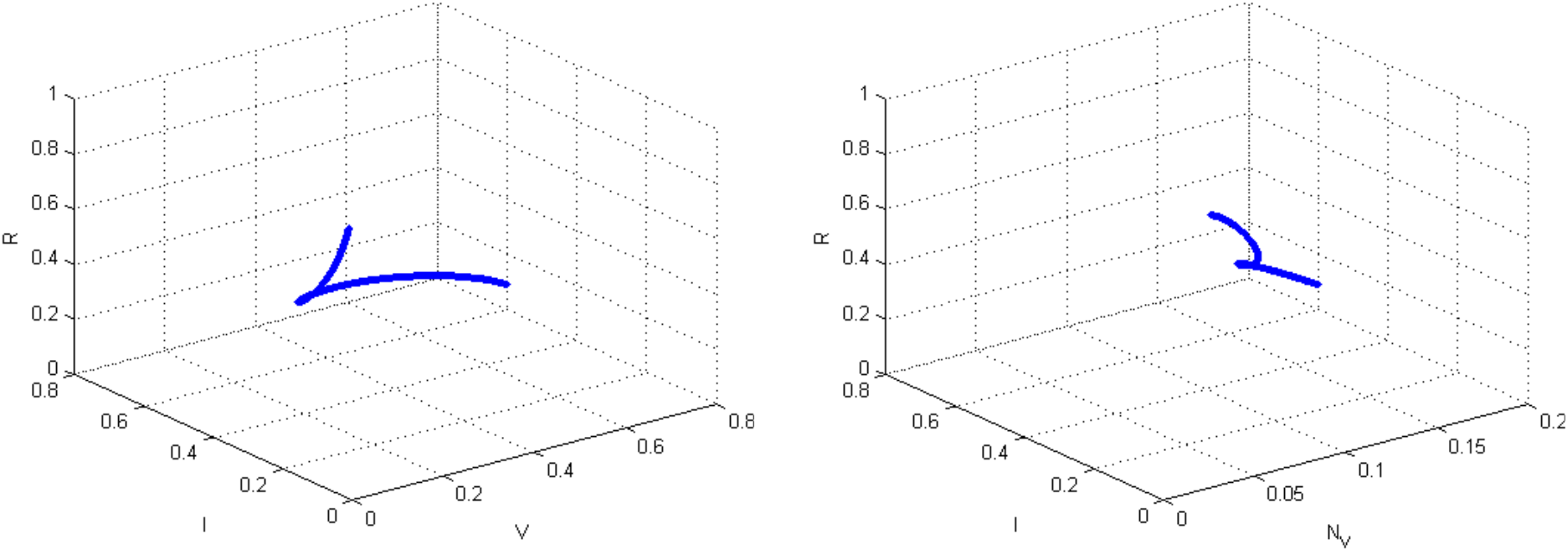
Cyclic behaviour of recovered individuals.

Figure 6 indicates the cyclic behaviour of recovered individuals. The first graph shows vaccinated individuals while the other displays non-vaccinated individuals. The graphs show that the cycle of vaccinated individuals is larger than non-vaccinated individuals. Furthermore, vaccinated individuals have greater time to reinfection when compared to that of non-vaccinated individuals.

**Figure 7:**
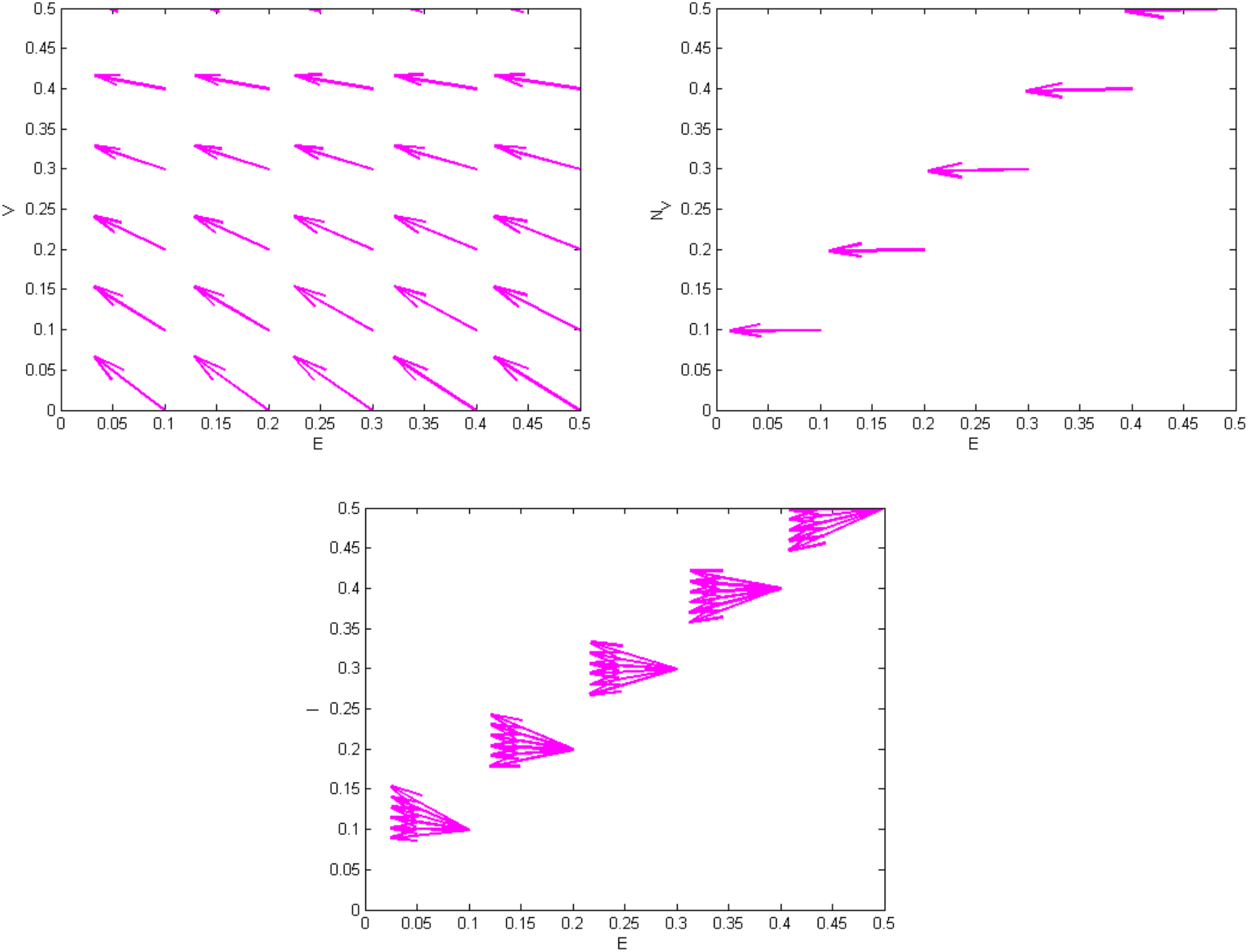
Behaviour of exposed individuals.

Figure 7 compares the behaviour of exposed individuals with vaccinated individuals, non-vaccinated individuals, and infected individuals, respectively. The rate of exposure is consecutive for non-vaccinated individuals when compared to vaccinated individuals. Furthermore, the transmission from exposed to infected is quite quick i.e. almost every individual who is exposed will get infected.

**Figure 8:**
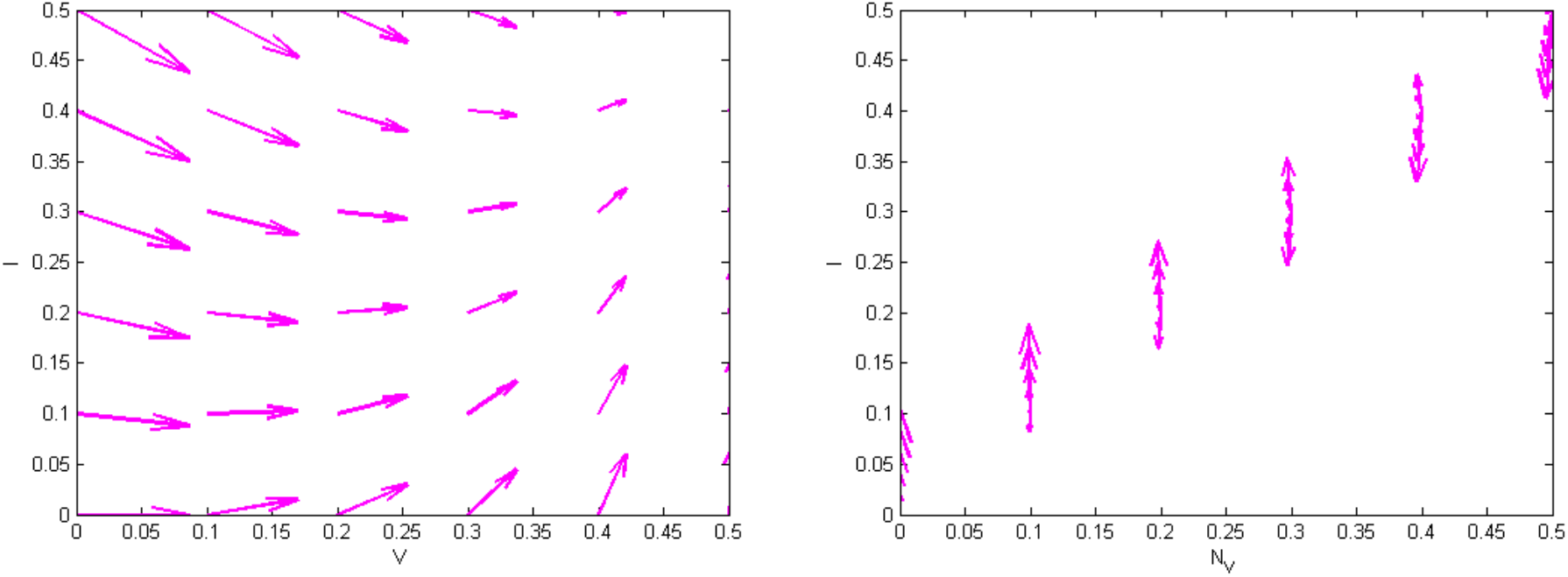
Behaviour of vaccinated and non-vaccinated individuals.

Figure 8 indicates that vaccinated individuals are infected gradually while non-vaccinated individuals are infection quickly. Consequently, non-vaccinated individuals are at higher risk of becoming infected with COVID-19.

**Figure 9:**
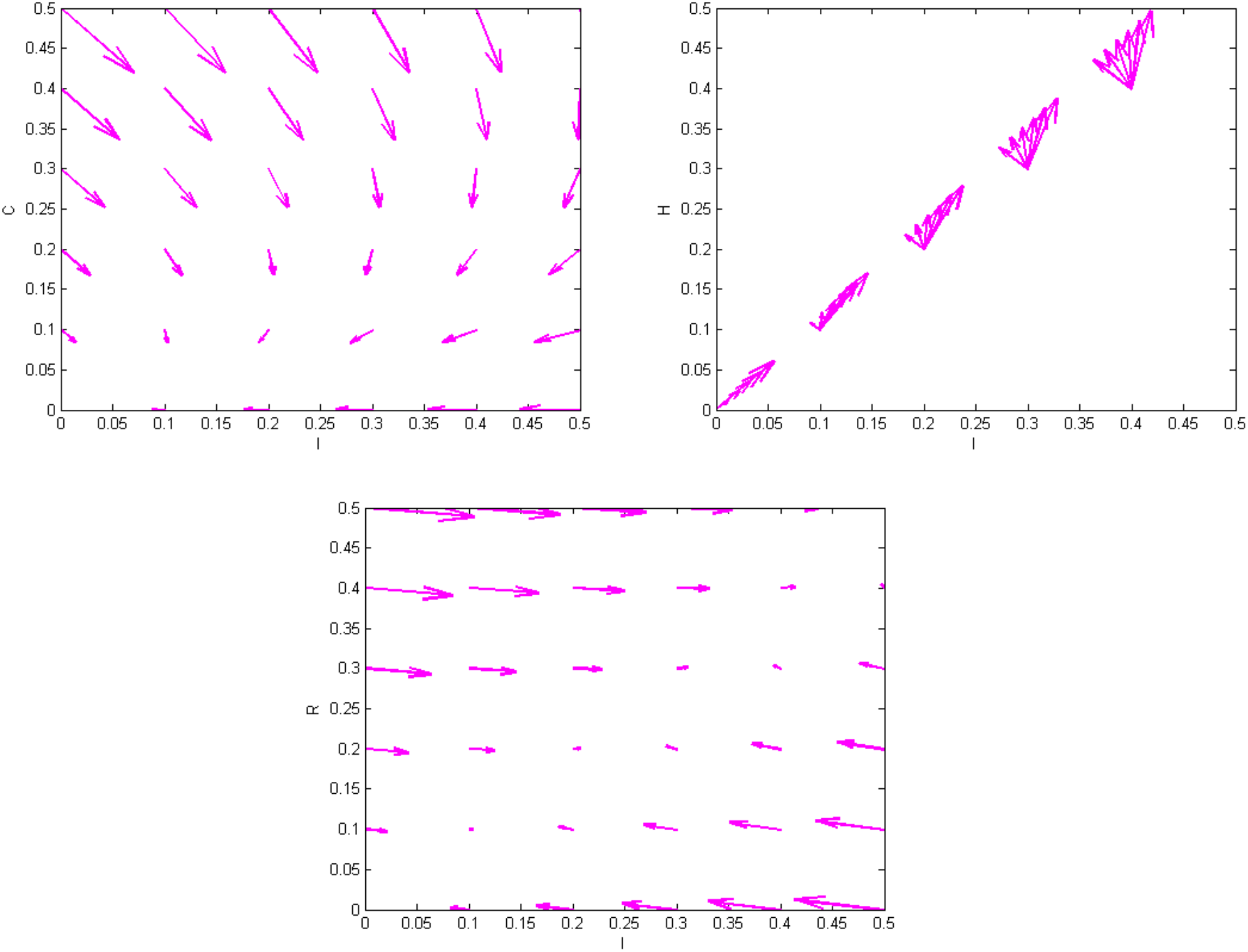
Behaviour of infected individual.

Figure 9 shows the behaviour of infected individuals towards critical, hospitalised and recovered individuals, respectively. If individuals are vaccinated early, the chances of becoming critically infection are low. However, this also indicates that infected individuals must receive treatment in hospitals before the infection progresses to critical stage. This is further elaborated by the last graph, which indicates that the chances of recovering by staying at home are negligible.

**Figure 10:**
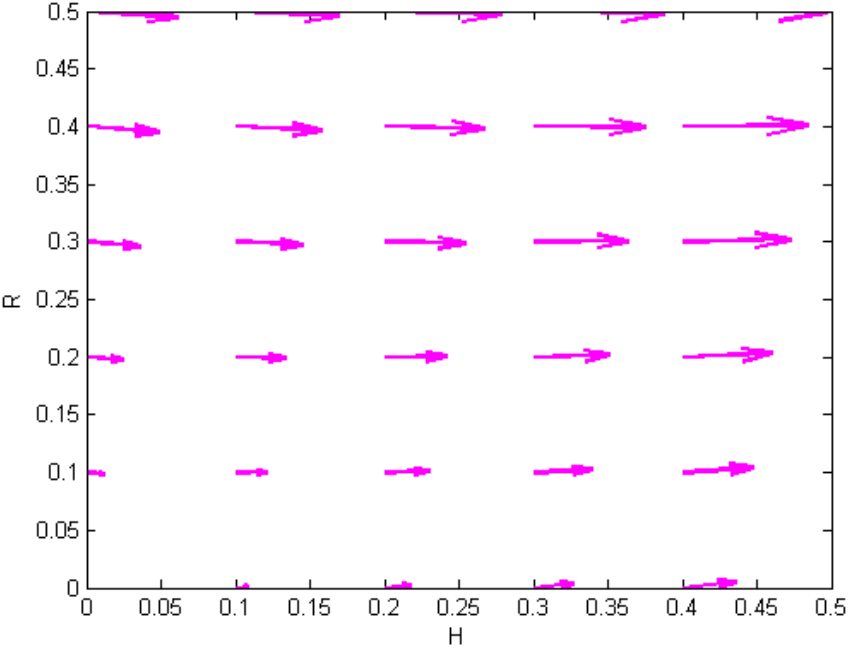
Behaviour of hospitalised individuals.

Figure 10 shows the behaviour of hospitalised individuals and their recovery. This figure further emphasizes that individuals recover only after proper hospitalization is received.

## 7. Conclusion

Our model indicates that a universal recommendation of BCG vaccination is beneficial in reducing the incidence rate of COVID-19. Moreover, the vaccination reduces the severity of the disease by reducing the hospitalization time for infected individuals. It also contributed to a higher rate of recovery for vaccinated individuals when compared to non-vaccinated individuals. The risk of reinfection is also reduced in vaccinated populations. Finally, the overall disease specific mortality of infected individuals is also reduced in vaccinated populations. Consequently, we validate the effectiveness of universal BCG vaccination policy in increasing herd immunity against COVID-19.

Considering that the BCG vaccination is established in preventing acute respiratory tract infections even in the elderly[13], vulnerable populations may be immunized with BCG vaccines until a COIVD-19 specific vaccine is developed. This strategy would be especially beneficial in the new-borns, elderly, and frontline essential workers. This study validates the call for clinical trials of BCG vaccine against COVID-19. Given that our model shows a significant reduction in incidence of COVID-19 in countries with universal BCG vaccination, we recommend all countries to conduct more clinical trials to establish and consider policy regarding BCG vaccinations for its citizens.

Our study may be subject to a few limitations. The study does not take into account confounding variables such as the potential differences in geographical and biological factors such as temperature, humidity, life expectancy, average income, social-cultural norms, ethnical genetic background, and mitigation between developing and developed countries. Due to these confounding variables, we cannot establish causality. The epidemiological data comparing vaccinated countries and non-vaccinated countries must be monitored throughout this pandemic to establish a causal impact in reducing incidence and mortality. Overall, our study serves to establish that individuals with BCG vaccinations have lower risk of getting COVID-19 infection, shorter hospital stays, and increased rate of recovery. Furthermore, countries with long-standing universal BCG vaccination policies have reduced incidence, mortality, and severity of COVID-19. Further research will focus on exploring the immediate benefits of vaccination to healthcare workers and patients as well as benefits of BCG re-vaccination. The results of this study will serve to inform and encourage further research and, ultimately, the creation of policy regarding universal BCG vaccination.

## Data Availability

Our conclusion is based on the data derived from our above mathematical calculations which are available to readers.

## Conflicts of Interest

The authors declare that we have no competing financial interests or personal relationships that could have appeared to influence the work reported in this paper.

## Acknowledgement

Second author (AHS) is funded by a Junior Research Fellowship from the Council of Scientific & Industrial Research (file no.-09/070(0061)/2019-EMR-I) and First three authors are thankful to DST-FIST file # MSI-097 for technical support to the Department of Mathematics, Gujarat University.

## Notes

### Competing Interest Statement

The authors have declared no competing interest.

## References

[1] J. Cohen, D. Normile, New SARS-like virus in China triggers alarm, Science (80). (2020). https://doi.org/10.1126/science.367.6475.234.

[2] M.L. Holshue, C. DeBolt, S. Lindquist, K.H. Lofy, J. Wiesman, H. Bruce, C. Spitters, K. Ericson, S. Wilkerson, A. Tural, G. Diaz, A. Cohn, L.A. Fox, A. Patel, S.I. Gerber, L. Kim, S. Tong, X. Lu, S. Lindstrom, M.A. Pallansch, W.C. Weldon, H.M. Biggs, T.M. Uyeki, S.K. Pillai, First case of 2019 novel coronavirus in the United States, N. Engl. J. Med. (2020). https://doi.org/10.1056/NEJMoa2001191.

[3] World Population Clock: Worldometer, (2020). https://www.worldometers.info/world-population (accessed March 29, 2020).

[4] Situation update worldwide, as of 21 April 2020, Eur. Cent. Dis. Prev. Control. (2020).

[5] M. Gursel, I. Gursel, Is Global BCG Vaccination Coverage Relevant To The Progression Of SARS-CoV-2 Pandemic?, Med. Hypotheses. (2020). https://doi.org/10.1016/j.mehy.2020.109707.

[6] A. Aaron Miller, Mac Josh Reandelar, Kimberly Fasciglione, Violeta Roumenova, Yan Li, G.H. Otazu, Correlation between universal BCG vaccination policy and reduced morbidity and mortality for COVID-19: an epidemiological study Aaron, J. Chem. Inf. Model. (2013). https://doi.org/10.1017/CBO9781107415324.004.

[7] A. Zwerling, M.A. Behr, A. Verma, T.F. Brewer, D. Menzies, M. Pai, The BCG world atlas: A database of global BCG vaccination policies and practices, PLoS Med. (2011). https://doi.org/10.1371/journal.pmed.1001012.

[8] J. Kleinnijenhuis, J. Quintin, F. Preijers, C.S. Benn, L.A.B. Joosten, C. Jacobs, J. Van Loenhout, R.J. Xavier, P. Aaby, J.W.M. Van Der Meer, R. Van Crevel, M.G. Netea, Long-lasting effects of bcg vaccination on both heterologous th1/th17 responses and innate trained immunity, J. Innate Immun. (2014). https://doi.org/10.1159/000355628.

[9] M.G. Netea, J. Quintin, J.W.M. Van Der Meer, Trained immunity: A memory for innate host defense, Cell Host Microbe. (2011). https://doi.org/10.10167j.chom.2011.04.006.

[10] S.J.C.F.M. Moorlag, R.J.W. Arts, R. van Crevel, M.G. Netea, Non-specific effects of BCG vaccine on viral infections, Clin. Microbiol. Infect. (2019). https://doi.org/10.1016/j.cmi.2019.04.020.

[11] J.T. Wu, K. Leung, G.M. Leung, Nowcasting and forecasting the potential domestic and international spread of the 2019-nCoV outbreak originating in Wuhan, China: a modelling study, Lancet. (2020). https://doi.org/10.1016/S0140-6736(20)30260-9.

[12] O. Diekmann, J.A.P. Heesterbeek, M.G. Roberts, The construction of next-generation matrices for compartmental epidemic models, J. R. Soc. Interface. (2010). https://doi.org/10.1098/rsif.2009.0386.

[13] R.J.W. Arts, S.J.C.F.M. Moorlag, B. Novakovic, Y. Li, S.Y. Wang, M. Oosting, V. Kumar, R.J. Xavier, C. Wijmenga, L.A.B. Joosten, C.B.E.M. Reusken, C.S. Benn, P. Aaby, M.P. Koopmans, H.G. Stunnenberg, R. van Crevel, M.G. Netea, BCG Vaccination Protects against Experimental Viral Infection in Humans through the Induction of Cytokines Associated with Trained Immunity, Cell Host Microbe. (2018). https://doi.org/10.1016/j.chom.2017.12.010.

